# Long-read transcriptome analysis using IsoRanker for identifying pathogenic variants in Mendelian conditions

**DOI:** 10.1101/2025.11.07.25339764

**Authors:** Yong-Han Hank Cheng, Adriana E Sedeño-Cortés, Jane E Ranchalis, Katherine M Munson, Mitchell R Vollger, Elsa Balton, Casie A Genetti, Undiagnosed Diseases Network, Genomics Research to Elucidate the Genetics of Rare Diseases consortium, University of Washington Center for Rare Diseases Research, Monica H Wojcik, Alan H Beggs, Michael J Bamshad, Chia-Lin Wei, Katrina M Dipple, Runjun D Kumar, Elizabeth E Blue, Gail Jarvik, Jessica X Chong, Daniela M Witten, Anne O’Donnell-Luria, Andrew B Stergachis

## Abstract

Identifying pathogenic non-coding variants that contribute to Mendelian conditions remains challenging as the functional impact of these variants on gene function is often unknown. We present IsoRanker, a long-read transcriptome sequencing-based framework that prioritizes functionally relevant non-coding variants by detecting genes and novel isoforms with outlier expression, allelic imbalance, and/or nonsense-mediated decay (NMD). We generated paired cycloheximide-treated and untreated fibroblast transcriptomes from 31 individuals (3 individuals with known transcript-altering rare variants and 28 individuals with unsolved conditions) and linked transcripts to phased long-read genomes. IsoRanker successfully recovered known transcript alterations in this cohort and remained robust in subsampling analyses to cohorts of 11 individuals and ∼5 million full-length transcripts per individual. However, performance was dependent upon *de novo* isoform caller choice, particularly for NMD-sensitive and novel isoforms. Among 28 previously unsolved cases, IsoRanker deprioritized most fibroblast-expressed candidate splice site variants while nominating new leads. In one individual, IsoRanker prioritized *HARS1*, revealing biallelic non-coding variants that together produced a partial *HARS1* loss-of-function and informed targeted therapy in this individual using histidine supplementation. These findings establish long-read, NMD-aware transcriptomics with IsoRanker as an effective approach for generating isoform-level functional evidence, improving classification of non-coding variants and supporting the diagnosis of individuals with rare diseases.

## Introduction

Identifying pathogenic variants underlying Mendelian conditions remains a significant challenge, particularly when variants reside in non-coding regions of the genome. While genome and exome sequencing can efficiently detect millions of variants in an individual, distinguishing pathogenic changes from benign variation can be difficult without functional context.

Non-coding variants are a significant contributor to Mendelian disease because they can disrupt gene function at multiple regulatory levels. For example, variants in non-coding regions such as splice donor sites, splice acceptor sites, branch points, and splicing regulatory motifs (e.g., exonic/intronic splicing enhancers and silencers) can produce transcripts that are degraded by nonsense-mediated decay (NMD) or translated into truncated or altered protein sequences with altered stability/localization. Importantly, this class of variation accounts for approximately one third of all pathogenic variants, with many of these variants causing disease via the creation of premature termination codons (PTCs) that result in NMD, underscoring the critical role of transcript alterations in Mendelian conditions.^1–3^

Combining genomic sequencing with short-read RNA sequencing (RNA-seq) has been established as a fruitful clinical approach for identifying transcript-altering pathogenic variants leading to alterations in expression or splicing patterns.^4^ Across a variety of tissues and conditions, this approach has been reported to increase diagnostic yield by 2%–24% vs DNA sequencing alone when evaluating cases of suspected Mendelian conditions.^5–9^ However, this approach is constrained by the limitations of short-read sequencing, which prevents comprehensive resolution of full-length transcript isoforms and fusion events. Additionally, short-read RNA outlier detection methods typically require hundreds of control samples to generate a robust baseline distribution for identifying outliers, which can be logistically challenging. Moreover, standard short-read RNA outlier detection methods only indirectly detect whether a transcript is subjected to NMD. As a result, pathogenic variants that act by promoting degradation of altered transcripts may go undetected, or require deep sequencing to detect, despite NMD being a common mechanism by which variants cause disease.^1^

Long-read full-length transcript sequencing offers the potential to overcome many of the limitations inherent to these short-read RNA-seq based outlier approaches. Specifically, long-read transcript data permits the *de novo* assessment of full-length isoforms, uncovering novel disease-associated isoforms in a haplotype-specific manner, including fusion transcripts and complex splicing events that are challenging to resolve using fragmented short-read data. However, despite the potential for long-read transcript sequencing to aid in the diagnosis of rare diseases, existing statistical tools for outlier analysis are not designed to leverage the unique advantages of long-read sequencing data.

To address this, we developed IsoRanker, a computational framework designed for per-individual prioritization of candidate genes and isoforms. IsoRanker takes in isoform-level expression data from multiple individuals and integrates these expression data to evaluate clinically relevant functional transcript-altering effects (i.e., NMD susceptibility, loss-of-expression, gain-of-expression, and allelic imbalance) (**Fig. 1**). Furthermore, IsoRanker was designed to handle paired long-read transcript data from both untreated samples and those treated with cycloheximide (CHX), an NMD inhibitor. This enables analysis of steady-state transcripts alongside transcripts stabilized by NMD inhibition. We demonstrate that this paired experimental design enables the discovery of pathogenic variants using samples with modest sequencing depth, thereby improving the cost associated with acquiring long-read transcript data. Importantly, using positive control samples we demonstrate that IsoRanker can accurately prioritize pathogenic variants using paired long-read transcript data from cohorts as few as 11 individuals. Finally, we demonstrated the application of IsoRanker to long-read transcript data from 28 individuals with suspected Mendelian conditions, showing the clinical utility of long-read data both in resolving the molecular basis of disease, as well as in ruling out variants that were falsely prioritized using short-read data. Overall, IsoRanker represents an advancement in the interpretation of non-coding variants. By leveraging the power of long-read transcript sequencing and providing a framework for functional analysis, IsoRanker enhances the diagnostic yield and allows for a more comprehensive understanding of the genetic basis of Mendelian conditions.

**Figure 1.**
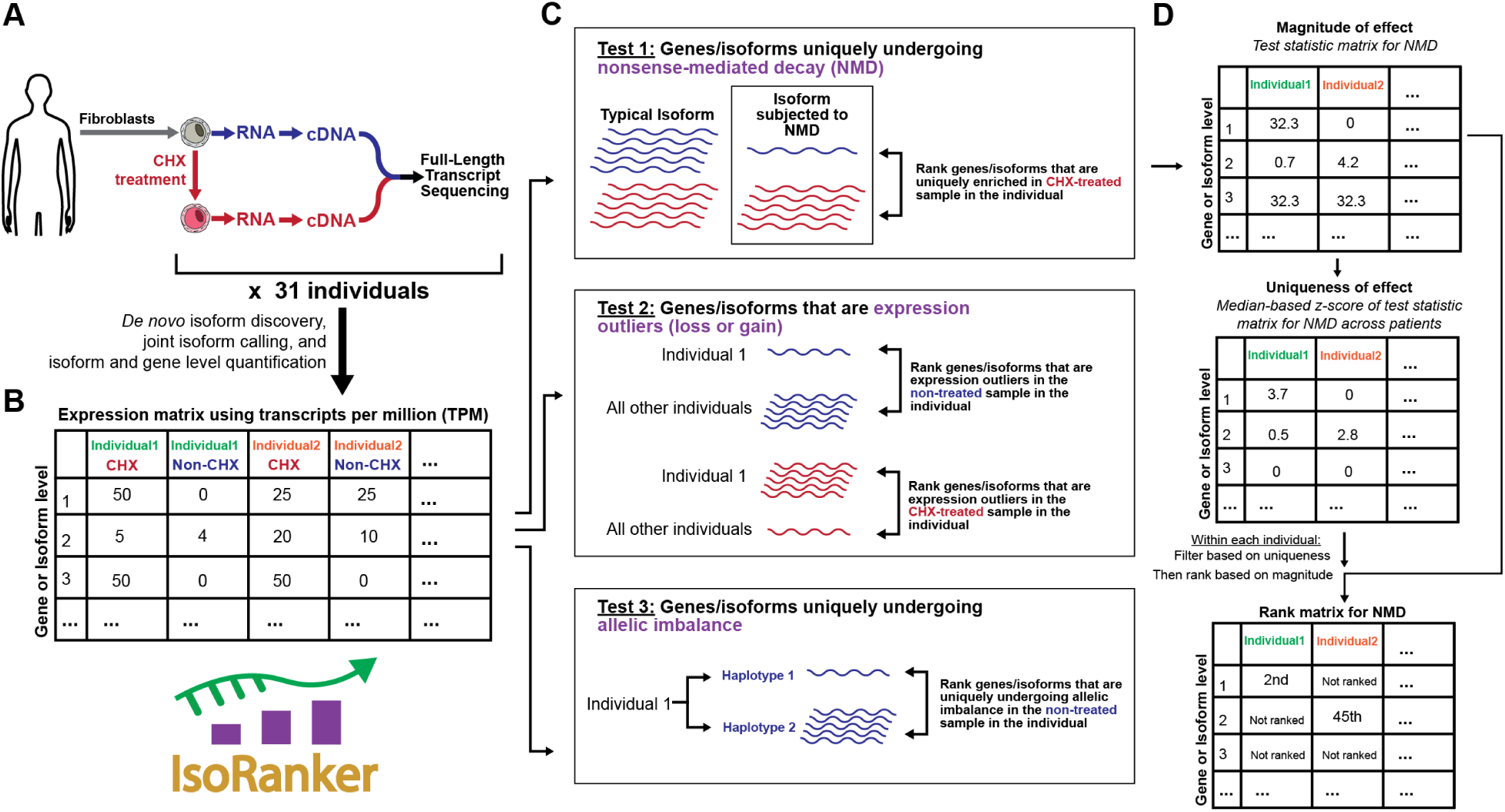
Overview of the IsoRanker workflow for prioritizing genes and isoforms with potential functional relevance in individual fibroblasts: (A) Fibroblasts from 31 individuals (28 unsolved plus 3 solved) are cultured with and without cycloheximide (CHX) treatment to inhibit nonsense-mediated decay (NMD). RNA is extracted, converted to cDNA, and subjected to full-length transcript sequencing for *de novo* isoform discovery, joint isoform calling, and quantification at the isoform and gene level. (B) An expression matrix (transcripts per million, TPM) is generated for CHX-treated and non-treated samples across all individuals. (C) IsoRanker performs three tests: (Test 1) Identification of genes/isoforms uniquely undergoing NMD, ranked by enrichment in CHX-treated samples. (Test 2) Detection of expression outliers (loss or gain) relative to other individuals, ranked separately for non-treated and CHX-treated samples. (Test 3) Identification of genes/isoforms uniquely undergoing allelic imbalance. (D) For each isoform/gene, we first calculated a test statistic that quantifies the magnitude of the functional effect observed. Using these test statistic values across all samples, we then computed a median-based z-score to assess whether the observed functional effect for a given isoform/gene in our individual was significantly different from that seen in other individuals. Finally, we retained isoforms/genes with z-scores above the 99.5^th^ percentile within the individual and subsequently ranked them within each individual according to their test statistic values to prioritize isoforms exhibiting the most pronounced functional alterations. Results from each test are stored in rank matrices for cross-individual comparisons.

## Material and Methods

### Study cohort

This study was approved by the National Institutes of Health Institutional Review Board (IRB) (IRB protocol #15HG0130) and the Boston Children’s Hospital IRB under protocol 10-02-0053, and written informed consent was obtained from all participants in the study. Individuals were enrolled from the Undiagnosed Diseases Network (UDN) and Boston Children’s Hospital Manton Center Gene Discovery Core [**Supplementary Table 4**] based on standard inclusion criteria, which required objective clinical findings relevant to their phenotype and the absence of a diagnosis despite comprehensive evaluation by healthcare providers.

### Identification of genetic variants

Short-read exome and genome sequencing was performed through the UDN (Baylor College of Medicine) or the Broad CMG. Long-read genome sequencing was performed with PacBio HiFi sequencing as previously described using the Fiber-seq protocol.^10^ Data were mapped to GRCh38. Variant calling was performed with DeepVariant^11^ (v1.5.0) for SNVs and indels and pbsv for SVs (https://github.com/PacificBiosciences/pbsv).

### Tissue culture

Individual-derived fibroblast cells were obtained via skin punch biopsies. Fibroblasts were cultured in DMEM (Gibco) supplemented with 10% FBS and 1% penicillin/streptomycin at 37°C in 5% CO2. We selected fibroblasts as the tissue source for transcriptomic analysis because, among clinically accessible tissues, fibroblasts offer the greatest consistency in gene expression profiles and demonstrate substantially higher expression levels across known disease-associated genes.^12,13^ This makes fibroblasts an ideal model for transcriptome-based diagnostics in rare Mendelian conditions.

### Evaluation of transcripts subjected to NMD

RNA from individual-derived fibroblast cells was separately isolated from untreated cells, as well as cells treated with cycloheximide (100 μg/mL, Sigma-Aldrich) for 6 hours before RNA extraction.

### RNA preparation and sequencing

Bulk mRNA was isolated using the Qiagen RNeasy Mini Kit (74104). RNA was subjected to polyA selection and cDNA synthesis as described in the standard PacBio Iso-Seq protocol, with the modification of using new cDNA amplification primers that allow for sample barcoding combined with MAS-seq directed concatenation primer sequences^14^ to enable subsequent MAS-seq concatenation. Following cDNA amplification, samples were pooled and concatenated. Individuals 2, 3, 5, 6, 13, 27, 28, 29, and 30 were pooled 2-plex, untreated and cycloheximide-treated cDNA from the same cell line, and sequencing libraries generated by adapting the MAS-seq for 10x Single Cell 3’ protocol to an 8-mer array (PacBio, 102-659-600). The remaining individuals were pooled together and the concatemerized sequencing library was generated with the Kinnex Full-Length RNA kit (PacBio, 103-072-000). Libraries were sequenced on the Revio platform on SMRT Cells 25M with Revio Chemistry V1 (PacBio, 102-817-900) with Adaptive Loading and 30-hour movies.

Each individual was sequenced to a depth of approximately 5 million reads per condition (cycloheximide-treated and untreated), yielding a total of approximately 10 million reads per individual. In cases where a sample exceeded this depth, reads were downsampled to achieve comparable coverage across samples. Specifically, full-length non-chimeric (FLNC) BAM files with more than 7 million reads were probabilistically downsampled to a target of ∼5 million reads. For each sample with more than 7 million reads, we computed a sampling probability p=5,000,000/N, where N is the total number of reads, and applied samtools view -s <seed.p> -b to retain each alignment independently with probability p using a fixed pseudorandom seed for reproducibility. The probability was specified to six decimal places to minimize rounding error in the target proportion. Because this procedure samples reads independently, the retained count follows a binomial distribution with mean Np≈5,000,000, thus the final counts are close to, but not exactly, the target. Samples at or below 7 million reads were not downsampled. All downstream analyses used the resulting downsampled BAM files. Data were processed using the PacBio IsoSeq v4.3.0 pipeline (https://isoseq.how/), aligned to GRCh38, and visualized using IGV.

### *De novo* isoform calling and quantification

Full-length non-chimeric (FLNC) reads were identified and clustered into high-quality consensus *de novo* isoforms using the IsoSeq v4.3.0 pipeline. The clustered consensus isoforms were aligned to the GRCh38 reference genome using pbmm2. To ensure consistent isoform naming across all samples, we merged the BAM files containing clustered FLNC reads from all samples using samtools merge, generating a single coordinate-sorted BAM file representing the combined set of clustered reads. This merged file was used for downstream processing to produce a unified set of non-redundant isoform models, ensuring that each isoform identifier (e.g., PB.1.1) corresponded to the same isoform model across all samples. Redundant isoforms were removed using isoseq collapse, producing a non-redundant set of unique transcript isoforms. Isoforms were classified as known or novel based on comparison to GENCODE v47 using Pigeon v1.4.0. Isoform quantification was performed by counting the number of FLNC reads supporting each isoform, as reported in the isoseq collapse and pigeon output files. All IsoSeq isoform calling, classification, quantification, and integration with phased variant data were performed using a standardized Snakemake workflow (IsoSeq_smk) available at https://github.com/StergachisLab/IsoSeq_smk.

In addition to the IsoSeq pipeline, we also performed *de novo* isoform calling and quantification using LRAA v0.3.0 (https://github.com/MethodsDev/LongReadAlignmentAssembler), IsoQuant^15^ v3.7.0, and TALON^16^ v5.0 to compare performance across independent long-read transcriptome analysis methods. For each tool, FLNC BAMs aligned to the GRCh38 reference genome were used as input. Isoform calling and quantification were performed according to each tool’s standard workflow.

### Phasing transcript reads

To assign transcript reads to haplotypes, we first performed genome sequencing using PacBio HiFi, as described above. SNV calls were generated with DeepVariant. We then used HiPhase^17^ v1.2.1 to produce phased variant calls across the genome. These phased variants were subsequently used within the <u>IsoSeq</u>_smk workflow in conjunction with WhatsHap^18^ v2.3 to assign aligned transcript reads to their respective haplotypes. When parental sequencing data was available, we applied a trio-aware phasing approach using a custom pipeline (https://github.com/mrvollger/k-mer-variant-phasing v0.0.1). This method uses SNVs identified by DeepVariant, then applies HiPhase to bin reads into phase blocks. The blocks are then assigned to the maternal or paternal haplotype by incorporating parental short-read genome data and running the meryl-based trio k-mer phaser (https://github.com/marbl/meryl).

### Expression matrix

An expression matrix was constructed for all 31 individuals, where rows represent isoforms and columns represent individual samples and values represent raw counts. This matrix was generated from IsoSeq’s read-level summaries and sample metadata, integrating haplotype assignments and treatment conditions (cycloheximide-treated or untreated). We similarly generated expression matrices separately using results from LRAA, IsoQuant, and TALON.

### Test statistic

All test statistic calculations were performed using IsoRanker (https://github.com/yhhc2/IsoRanker) with the 31-individual expression matrix. Prior to test statistic calculations, raw counts were converted to transcripts per million (TPM). We applied a suite of functional effect-specific scoring metrics (test statistics) for each gene and isoform in each individual to quantify NMD, loss-of-expression, gain-of-expression, and allelic imbalance. We evaluated the following functional effects at both the gene and isoform levels, where applicable. Each functional effect captures a specific pattern of transcript alteration relevant to potential pathogenic mechanisms.

### Nonsense-Mediated Decay (NMD)

This functional effect detects isoforms/genes that are stabilized in the presence of cycloheximide, suggesting that the isoform/gene is normally targeted for degradation via nonsense-mediated decay (NMD). The test statistic captures the degree of increase in expression upon cycloheximide treatment, using a log-scaled ratio of cycloheximide-treated to untreated TPM values, adjusted by expression magnitude:

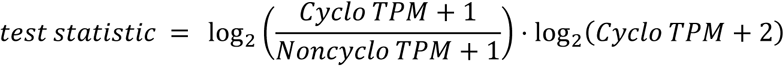

### NMD Rare Steady-State Transcript

This effect identifies genes with isoforms that are unusually rare in the untreated condition (steady-state) but present upon cycloheximide treatment, indicating potential NMD. The test statistic contrasts bin-wise transcript distributions:

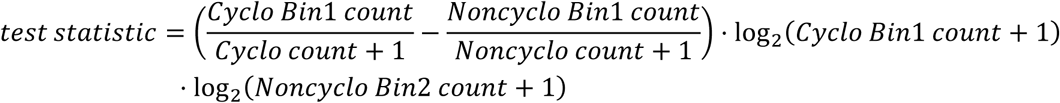

where Bin1 represents the set of isoforms with very low relative expression in the untreated sample. Specifically, these are isoforms whose read counts contribute less than 1% of the total read count for the gene in that sample (i.e., isoform proportion < 0.01 relative to the gene’s total expression). Bin2 represents isoforms with higher relative expression (proportion ≥ 0.01) in the untreated sample.

### Noncycloheximide Loss-of-Expression (Noncyclo LOE)

This effect identifies isoforms/genes showing unusually low expression in untreated conditions relative to other individuals. The test statistic quantifies the fold-reduction of expression in the individual compared to the minimum expression seen across other individuals, scaled by the median expression in other individuals:

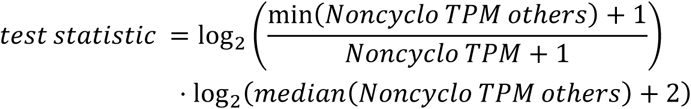

### Noncycloheximide Gain-of-Expression (Noncyclo GOE)

This effect identifies isoforms/genes showing abnormally high expression in untreated conditions. The test statistic measures the fold-increase of expression in the individual compared to the maximum expression in other individuals, scaled by the median expression of others:

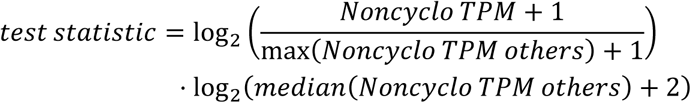

### Cycloheximide Gain-of-Expression (Cyclo GOE)

This effect identifies isoforms/genes showing outlier gain-of-expression in cycloheximide-treated samples. The test statistic compares the individual’s cycloheximide expression to the maximum of other individuals, similarly scaled by the median:

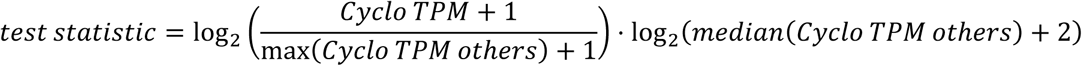

### Noncycloheximide Allelic Imbalance

This effect measures imbalance in allelic expression in untreated conditions. The test statistic quantifies deviation from equal haplotype expression using log-scaled haplotype ratios and overall haplotype read support:

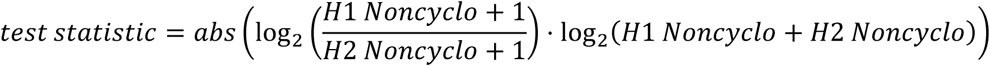

with filtering applied to require sufficient phased reads (≥10) and proportion of phased reads (≥10%).

The functional effects were evaluated at both the gene and isoform level. Specifically, at the gene level, we evaluated for: (a) Nonsense-mediated decay (NMD); (b) Noncycloheximide loss-of-expression (Noncyclo_LOE); (c) Noncycloheximide gain-of-expression (Noncyclo_GOE); (d) Cycloheximide gain-of-expression (Cyclo_GOE); (e) NMD rare steady-state transcript; and (f) Noncycloheximide allelic imbalance.

At the isoform level, we evaluate for: (a) Nonsense-mediated decay (NMD); (b) Noncycloheximide loss of expression (Noncyclo_LOE); (c) Noncycloheximide gain of expression (Noncyclo_GOE); (d) Cycloheximide gain of expression (Cyclo_GOE); and (e) Noncycloheximide allelic imbalance.

#### Outlier analysis and ranking

Outlier detection and ranking were conducted using IsoRanker. For each functional effect, we computed a test statistic using the predefined formulas tailored to the effect. A median-based z-score was then computed for each gene or isoform for each individual:

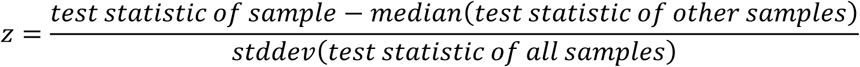

where the median excluded the index sample, and a z-score of 0 was assigned where standard deviation was zero.

Outlier detection was based on this z-score distribution within each individual. For each individual, the 99.5th percentile z-score was determined across all genes or isoforms evaluated. Only genes or isoforms with z-scores above this threshold were retained for further analysis and the rest are not ranked. For example, if an individual had 1,000 isoforms, the top 5 isoforms with the highest z-scores would pass this filter. The retained isoforms were then ranked within the individual based on the magnitude of their test statistic, independently for each functional effect. This approach ensured that outlier detection was both stringent and personalized, focusing on the most extreme deviations specific to each individual and functional category.

#### IsoRanker execution

The IsoRanker process was executed on a single CPU thread using the University of Washington’s Klone high-performance computing cluster, requiring approximately 4 hours and up to 280 GB of RAM to process an expression matrix generated from the 31 individuals. Full script can be found at https://github.com/yhhc2/IsoRanker/blob/main/examples/Expression_AllelicImbalance_NMD/Expression_AllelicImbalance_NMD.ipynb. IsoRanker results can be visualized in tabular format and volcano plots with IsoRanker_vis (https://github.com/StergachisLab/IsoRanker_viz), an R Shiny visualization extension that uses output from IsoRanker. All code used to generate IsoRanker figures and reproduce the analyses presented in this study is available at https://github.com/yhhc2/IsoRanker/tree/main/Paper.

## Results

### Establishing a rare disease long-read transcript dataset for disease discovery

To evaluate how long-read transcript data could be used in evaluating individuals with previously unsolved conditions, we first sought to generate a comprehensive dataset of long-read transcript data across 31 individuals. This dataset included 3 individuals with previously identified transcript-altering variants to serve as positive controls (i.e., a homozygous branch point variant, a heterozygous splice donor variant, and a promoter structural variant). In addition, this dataset included 28 individuals suspected of having an undiagnosed Mendelian condition despite non-diagnostic exome/genome sequencing, with 7 of the individuals having prior non-diagnostic short-read RNA sequencing with fibroblasts (**Table 1**). Given the unique clinical phenotypes of the individuals in this cohort, we assumed that no two individuals would likely have the same underlying genetic cause, allowing us to use the individuals as mutual controls in a cohort-based analysis framework. We subjected fibroblast cultures from each individual to long-read PacBio genomic DNA sequencing using Fiber-seq-treated DNA as well as long-read transcript sequencing using steady-state polyadenylated RNA (non-CHX). Furthermore, we also performed long-read transcript sequencing on polyadenylated RNA obtained after treating fibroblast cultures with an NMD inhibitor (CHX-treated).

**Table 1.**
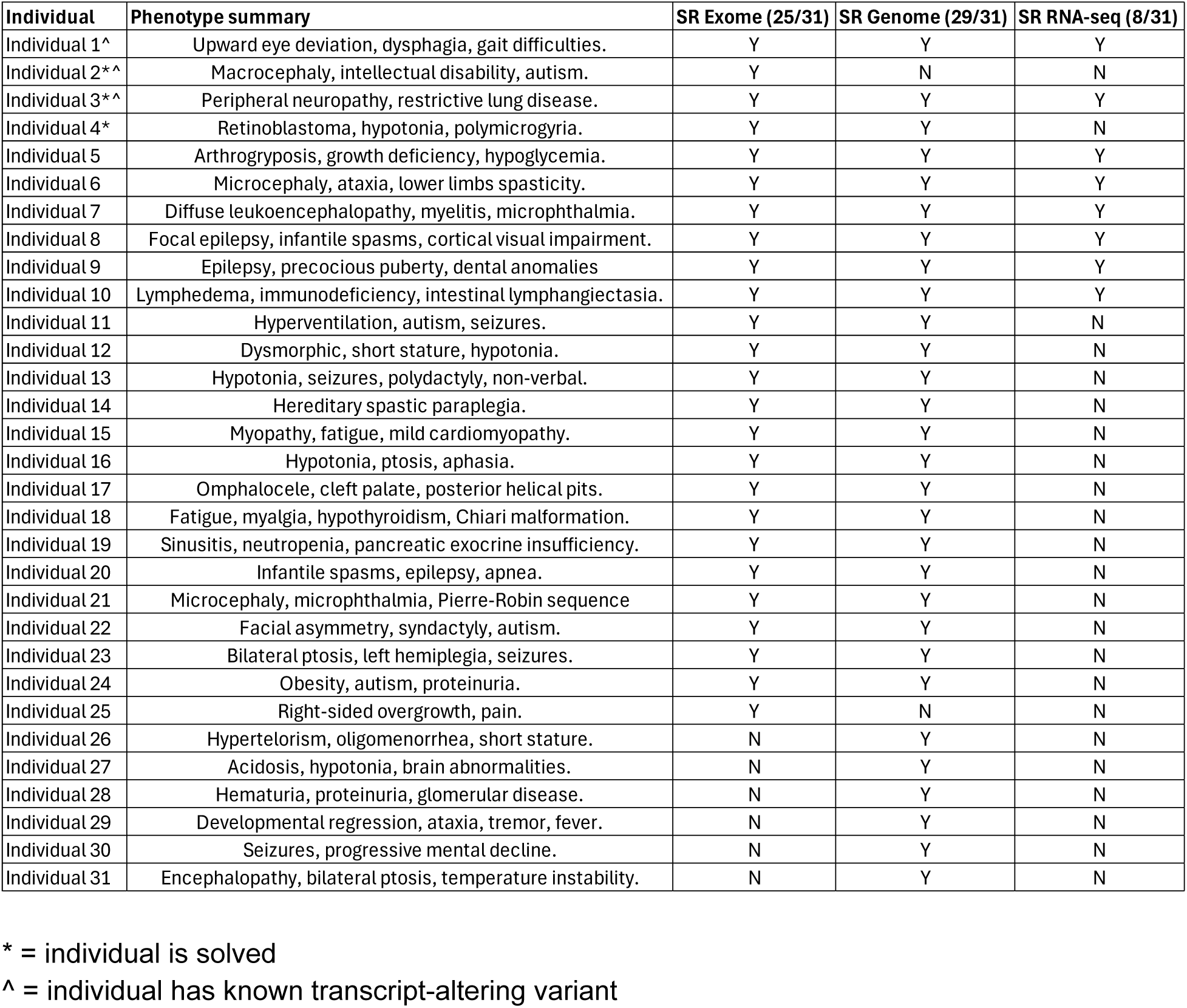
Phenotype summary of 31 individuals. * = individual is solved ^ = individual has known transcript-altering variant

Long-read PacBio genomic DNA sequencing on these samples yielded an N50 of ∼19,742 bp with a ∼47x mean coverage of hg38, permitting the phasing of ∼92.50% of heterozygous genomic variants within each individual, with 13 of these being parentally phased using parental short-read genome sequencing data. For the long-read transcript sequencing, two of the control samples were sequenced to ∼40 million full-length non-chimeric (FLNC) reads, with the rest of the samples being sequenced to a median of 6.2 million FLNC reads. The median N50 for transcript length was 2,134 bp for the non-CHX samples and 2,325 bp for the CHX-treated samples.

To quantify transcript features between samples, each full-length transcript sample was downsampled to ∼5 million FLNC reads, haplotype-phased using the paired genomic data, and then subjected to joint *de novo* isoform discovery using IsoSeq and subsequent classification using GENCODE v47. Overall, this resulted in a median of ∼38,481 unique genes identified per sample, including antisense genes and novel fusion genes, with on average 9 unique isoforms identified per gene in each individual (**Supplementary Table 2**). In the cohort, a total of 100,274 isoforms were discovered with 10 or more reads in any sample, and 4,579 of these isoforms were classified as fusion isoforms between multiple genes. Evaluation of the *SRSF6* poison exon^19^ in each sample confirmed robust NMD inhibition selectively in each CHX-treated sample, with hierarchical clustering of transcript abundances demonstrating that CHX treatment was the primary feature driving sample clustering (**Supplementary Fig. 1**). Furthermore, evaluation of known imprinted genes^20,21^ demonstrated appropriate haplotype phasing of each sample (**Supplementary Fig. 2**). Overall, this demonstrated that this experimental approach enabled us to robustly quantify the NMD-responsiveness and haplotype-selectivity of full-length transcripts across all 31 individuals.

### Benchmarking IsoRanker for prioritizing functional transcript-altering variants

Next, to identify specific genes or isoforms that are uniquely altered within a given individual, we first developed a statistical framework based on a set of test statistics that quantify distinct functional changes in an individual’s transcriptome (i.e., NMD susceptibility, loss-of-expression, gain-of-expression, and allelic imbalance). Importantly, these test-statistics are grounded in *de novo* long-read transcript isoform data, which permits the identification of altered isoforms absent from existing isoform catalogs. Next, to identify whether a functional change in the transcriptome is uniquely present within an individual, we applied a median-based Z-score method that permitted the ranking of all isoforms/genes from a given individual based on whether that particular isoform/gene was uniquely altered within that individual. Overall, this approach permits the identification of genes and isoforms that may harbor functionally relevant non-coding or coding variants, irrespective of whether that gene harbors a genetic variant that was prioritized based on orthogonal genetic sequencing analysis. We implemented this statistical framework in a Python package called IsoRanker (**Fig. 1**).

We benchmarked this transcriptome-wide ranking approach against three cases with previously established functional transcript alterations (**Fig. 2**). Specifically, Individual 1 harbors a heterozygous structural variant that impacts the promoter of *VTA1* that was shown using short-read RNA-seq of individual blood cells to cause loss-of-expression. IsoRanker ranked *VTA1* as the top hit genome-wide for uniquely being allelically imbalanced and within the top 20 genes genome-wide for uniquely having loss-of-expression in this sample (**Fig. 2a**), validating the ability of IsoRanker to detect and prioritize allelic imbalance and loss-of-expression via a transcriptome-wide search. While this confirmed the functional effect of the structural variant, inheritance from an unaffected parent suggested it was not disease-causing in this case, emphasizing the necessity of integrating functional data with segregation and phenotype. In addition, in Individual 3, IsoRanker appropriately ranked *MFN2* within the top 10 genes genome-wide having NMD sensitivity (**Fig. 2c**), consistent with this individual’s diagnosis of autosomal recessive Charcot-Marie-Tooth disease type 2A (CMT2A, MIM 617087), which was previously identified to be caused by a homozygous *MFN2* c.600-31T>G variant that disrupts a branchpoint.^22^ Finally, in Individual 2, IsoRanker appropriately ranked a *SET* isoform within the top 20 isoforms genome-wide for uniquely having gain-of-expression within this individual (**Fig. 2b**), consistent with this individual’s prior diagnosis of *SET*-related intellectual disability (MIM 618106) based on the genetic finding of a *de novo SET* c.663+5G>C splice donor variant. Our long-read data demonstrated that this variant likely causes *SET*-related intellectual disability via the generation of two aberrant isoforms: one with complete exon 6 skipping (in-frame), and another with partial exon 6 deletion (out-of-frame, leading to NMD).

**Figure 2.**
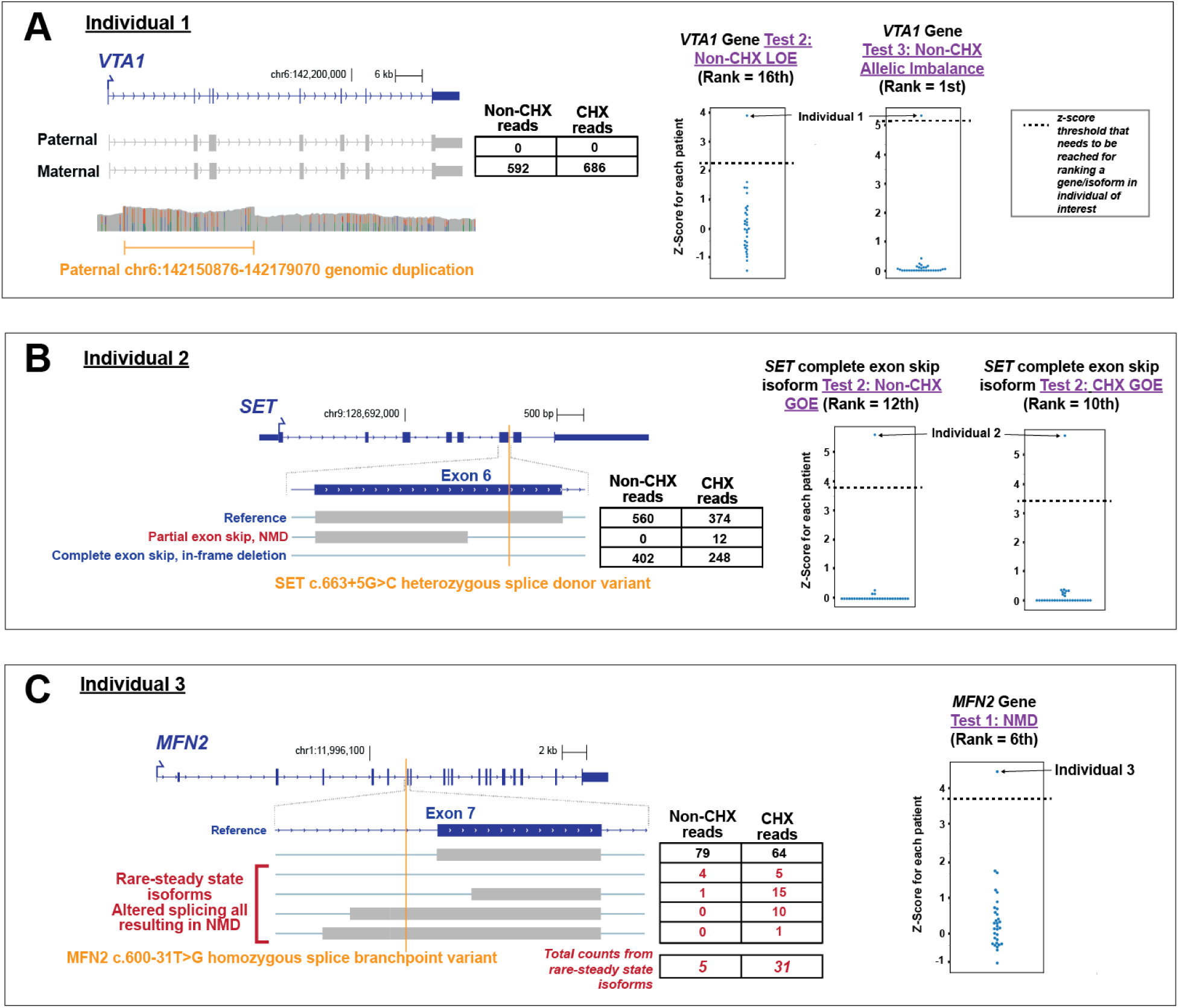
IsoRanker successfully detects five known transcript alterations: For each case, the genome browser view shows the reference isoform in blue and observed isoforms in gray, with the variant position marked by an orange vertical line. Tables list read counts in non-treated (Non-CHX) and CHX-treated fibroblasts. Right-hand plots display the median-based z-score of the test statistic for the indicated gene/isoform across all individuals, with the individual of interest labeled. (A) Individual 1 (*VTA1*): A paternal genomic duplication (chr6:142150876–142179070) results in complete loss of paternal allele expression for *VTA1*. Expression is detected exclusively from the maternal allele in both CHX and Non-CHX conditions. *VTA1* ranks 16th in the loss-of-expression (LOE) test and 1st in the allelic imbalance test for this individual. (B) Individual 2 (*SET*): A heterozygous splice donor variant (c.663+5G>C) in *SET* leads to partial exon skipping (NMD) and complete exon skipping (in-frame deletion). The *SET* isoform with in-frame deletion was ranked highly for gain-of-expression (GOE) regardless of CHX treatment for this individual. (C) Individual 3 (*MFN2*): A homozygous splice branchpoint variant (c.600-31T>G) in *MFN2* causes altered splicing that generates rare isoforms predicted to undergo nonsense-mediated decay (NMD). Rare steady-state isoforms are enriched after CHX treatment (5 Non-CHX vs. 31 CHX). The NMD test ranks *MFN2* 6th for this individual.

We next evaluated the cohort sample size and read depth required for IsoRanker to accurately rank these known transcript alterations (**Fig. 3**). We found that decreasing the sample size from 31 to 11 individuals resulted in a nearly identical prioritization of these known transcript alterations, but further reduction beyond 11 individuals diminished the performance of IsoRanker. In addition, downsampling the sequencing depth from 5 to 3 million FLNC resulted in a nearly identical prioritization of these known transcript alterations, but further reduction beyond 3 million FLNC diminished the performance of IsoRanker for these known transcript alterations that had moderate transcript expression levels in fibroblasts (**Supplementary Table 1**). Consequently, this indicates that IsoRanker is robust to cohorts that may include only a dozen individuals at a relatively economical sequencing depth (i.e., current PacBio sequencing typically results in 40-60 million FLNC from one SMRT cell).

**Figure 3.**
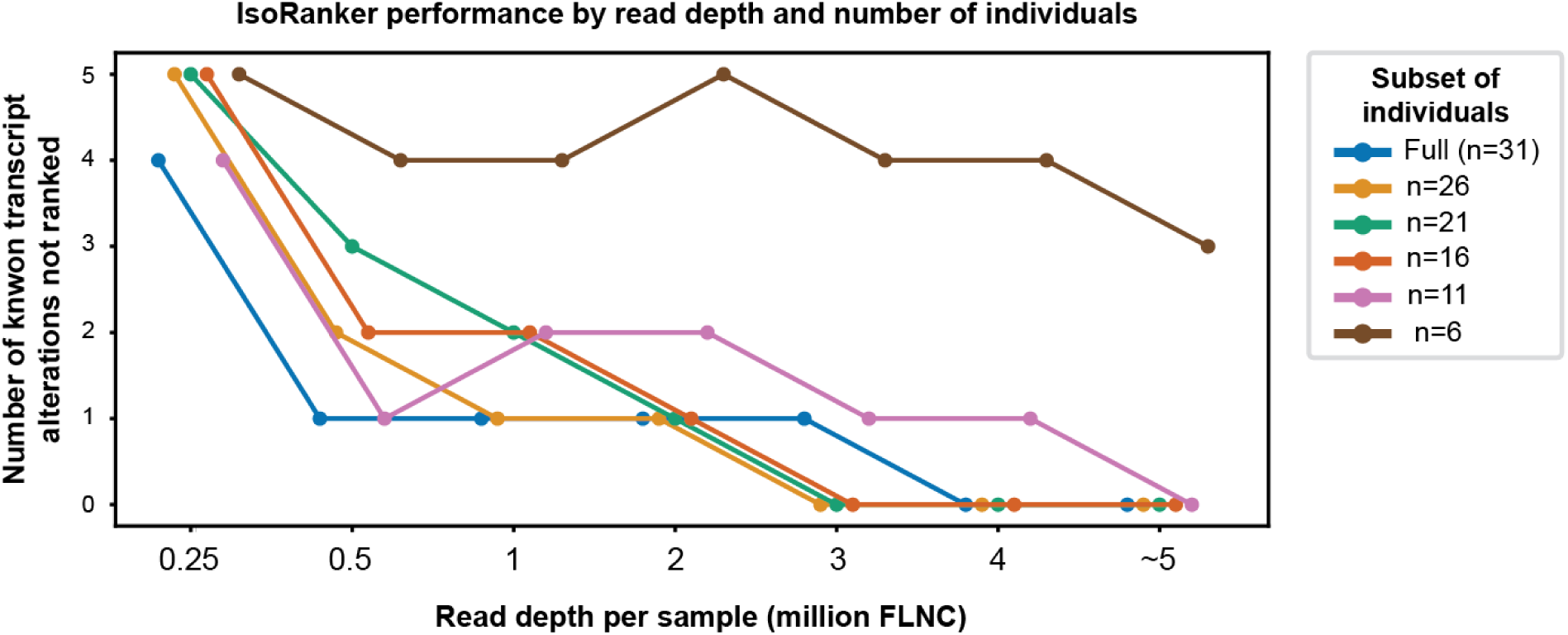
Assessing read counts and cohort sample sizes required to rank previously known transcript alterations: We used the five previously known transcript alteration events to evaluate the impact of read depth and cohort sample size on the ability to assign a rank to an event. The number of events not ranked generally decreases with increasing read depth, although smaller individual subsets (e.g., 6 individuals) consistently are unable to rank these known events.

### Impact of *de novo* isoform discovery on identifying transcript-altering variants

IsoRanker is grounded around *de novo* isoform discovery to identify novel isoforms that may be contributing to the pathogenesis of an individual’s disease. Multiple computational tools exist for performing *de novo* isoform discovery using long-read transcript data, and we next sought to evaluate the performance of these tools for rare disease diagnosis. To accomplish this, we performed *de novo* isoform discovery using four distinct computational tools (i.e., IsoSeq, IsoQuant, LRAA, TALON) and evaluated their performance using IsoRanker. We observed that these *de novo* isoform callers showed a large difference in the number of isoforms identified using the same input data, with IsoSeq (355,794 isoforms/sample) and TALON (343,977 isoforms/sample) identifying more isoforms compared to IsoQuant (48,809 isoforms/sample) and LRAA (90,027 isoforms/sample). This is likely attributed to differences in their preference for lumping versus splitting when calling new isoforms.

Application of IsoRanker to *de novo* isoforms identified using each of the four callers was still able to correctly prioritize the gene that was subjected to loss-of-expression and allelic imbalance (**Fig. 4a**). However, only IsoSeq and TALON were able to uniformly prioritize the isoform that had novel gain-of-expression and the gene that had rare isoforms subjected to NMD. On further evaluation of this, we identified that the other *de novo* isoform callers failed to assign the reads resulting from the pathogenic splice-altering variants to a novel isoform and instead lumped these reads with the reference isoform for the gene (**Fig. 4b**). Consequently, the predilection of a *de novo* isoform caller to either lump or split reads into unique isoforms appears to have a direct impact on the ability of that caller to identify novel isoforms relevant to rare diseases.

**Figure 4.**
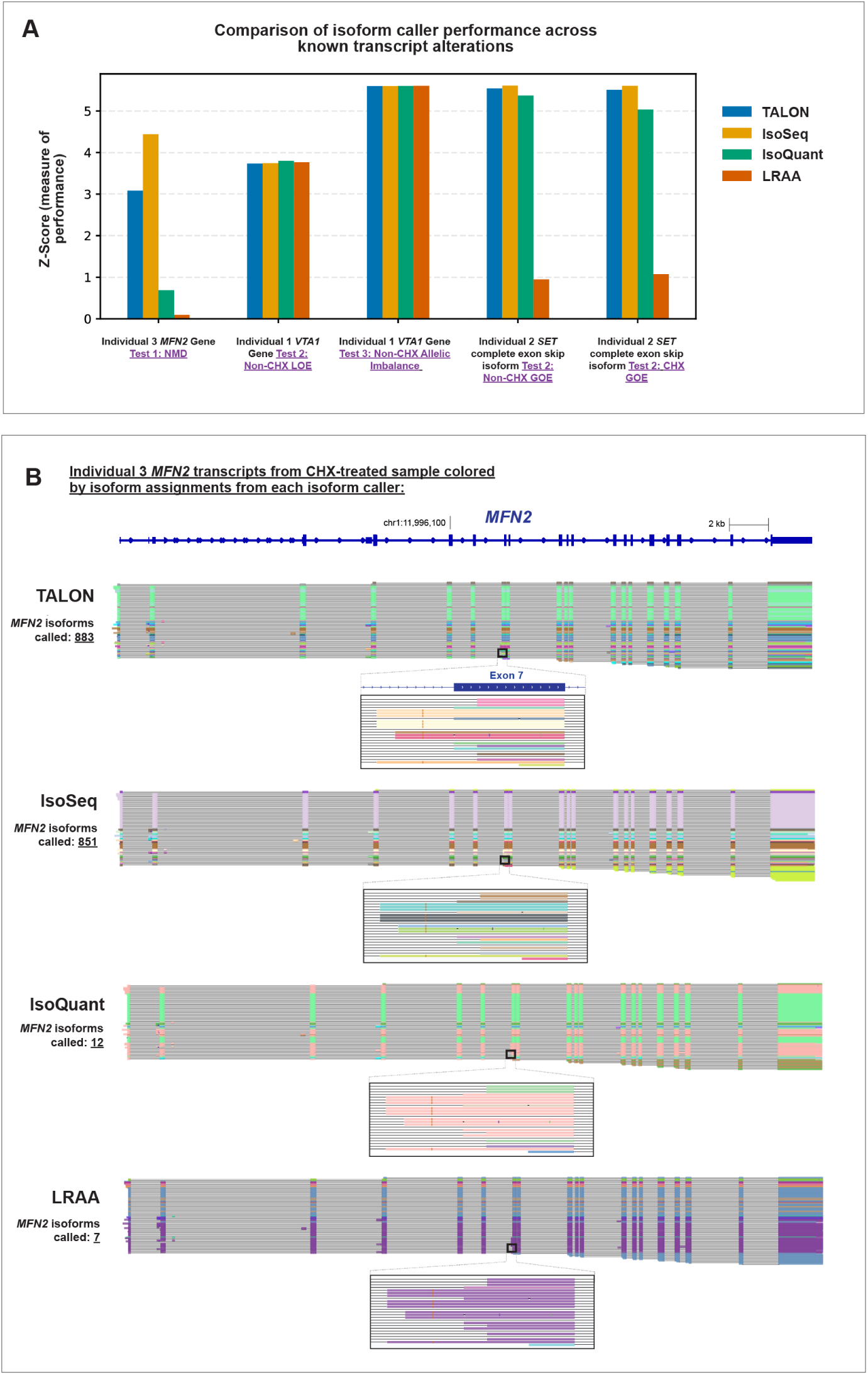
Comparison of different *de novo* isoform callers for identifying pathogenic transcripts: (A) We applied IsoRanker on expression matrices generated with four different isoform callers. We then evaluated the median-based z-score calculated from IsoRanker for the five previously known transcript alteration events. (B) Genome browser view of *MFN2* full-length transcripts in Individual 3 colored by isoform assignment from each isoform caller, illustrating TALON and IsoSeq as splitters and IsoQuant and LRAA as lumpers when calling isoforms.

### Application of IsoRanker for prioritizing candidate variants

After establishing IsoRanker’s ability to identify known transcript-altering variants, we evaluated whether IsoRanker could provide functional evidence for evaluating candidate transcript-altering variants. Specifically, based on the paired genome/exome data, 24 putative splice altering variants were prioritized, 10 of which (42%) localized to genes robustly expressed in fibroblasts (**Supplementary Table 1**). Evaluation of the transcript sequence data and the IsoRanker output for these 10 variants revealed that 8 of them had no functional impact on splicing within the individual fibroblast cells. For example, a *SEC31A* c.-174+1G>T splice donor variant that was previously prioritized based on the presence of an additional *SEC31A* c.3647A>G missense variant in trans, was found to actually localize to the gene *THAP9-AS1* based on the full-length transcript data, with the putative *SEC31A* isoform (ENST00000355196.6) being identified as a minimally expressed isoform in all tissues (**Supplementary Fig. 3**), indicating that this variant is likely not functionally altering *SEC31A* (**Fig. 5**). In contrast, we identified that two of the 10 variants did indeed impact splicing. Specifically, we identified that the *MTMR14* c.1769+1G>C variant resulted in the in-frame skipping of exon 18 of *MTMR14*, and the *SLC41A2* c.736-1G>A variant resulted in the out-of-frame skipping of exon 5 of *SLC41A2*. However, whereas the *MTMR14* functional effect was prioritized using IsoRanker, the *SLC41A2* functional effect was not, owing to the splitting of *SLC41A2* transcript reads across 22 isoforms using IsoSeq and diluting the signal for any single isoform, highlighting a potential drawback associated with *de novo* isoform callers over-splitting.

**Figure 5.**
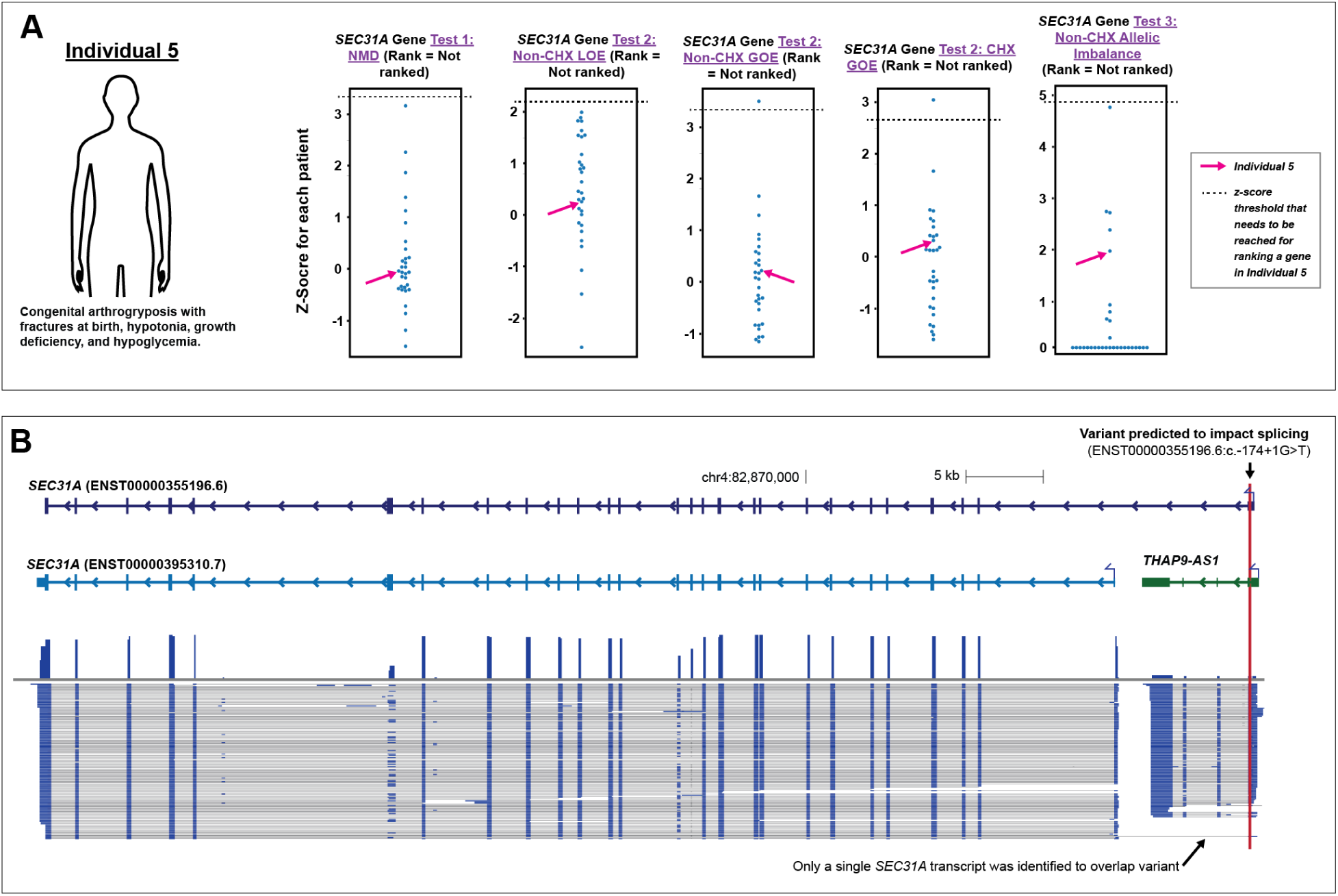
IsoRanker rules out candidate genetic cause: (A) Individual 5 has arthrogryposis multiplex congenita, fractures at birth, hypotonia, and growth hormone deficiency. Exome sequencing identified compound heterozygous *SEC31A* variants: a missense variant (c.3647A>G) and a splice donor variant (c.-174+1G>T). Z-score of the test statistic was plotted for *SEC31A* across all individuals for each of the gene-level tests. *SEC31A* was not ranked in any test. (B) Genome browser view showing *SEC31A* (two reference transcripts in light and dark blue) and the neighboring gene *THAP9-AS1* (green). Observed isoforms from individual fibroblasts are shown below, with the splice donor variant indicated by a red vertical line near the 3′ end of *THAP9-AS1*. Isoform data reveal that most *SEC31A* transcripts initiate downstream of the splice donor variant, preserving normal transcript structure. The affected isoform (NM_001318120.2) is minimally expressed, explaining the absence of functional alterations detected by IsoRanker and ruling out *SEC31A* as the disease gene.

### Diagnosis of a previously unsolved case using IsoRanker

Having benchmarked the performance of IsoRanker, we next sought to determine whether any prioritized genes may be diagnostic for the unsolved cases in our cohort. Specifically, in Individual 6, IsoRanker prioritized *HARS1* as a top candidate across multiple categories (**Fig. 6a**): NMD (ranked 4th), loss-of-expression (ranked 1st), and allelic imbalance (ranked 4th). Furthermore, IsoRanker prioritized the fusion gene *DND1-HARS1* as a top candidate for gain-of-expression (ranked 4th). Analysis of this individual’s paired genetic sequencing data revealed compound heterozygous rare non-coding variants in *HARS1*, including a maternally inherited c.951+5G>T splice donor variant and a paternally inherited c.*331A>T variant at the 3’ UTR (**Fig. 6b**). The long-read transcript data revealed that the c.951+5G>T variant causes exon 9 skipping, producing out-of-frame transcripts that are targeted by NMD. Furthermore, the long-read transcript data revealed that the c.*331A>T variant occurs at the first position of the primary polyadenylation signal (AAUAAA) for *HARS1*, leading to formation of a *DND1-HARS1* fusion isoform, also subject to NMD. Using ACMG/AMP criteria ^23,24^, the splice donor variant is classified as likely pathogenic and the 3’ UTR is classified as a variant of uncertain significance (VUS) (**Supplementary Table 3**).

**Figure 6.**
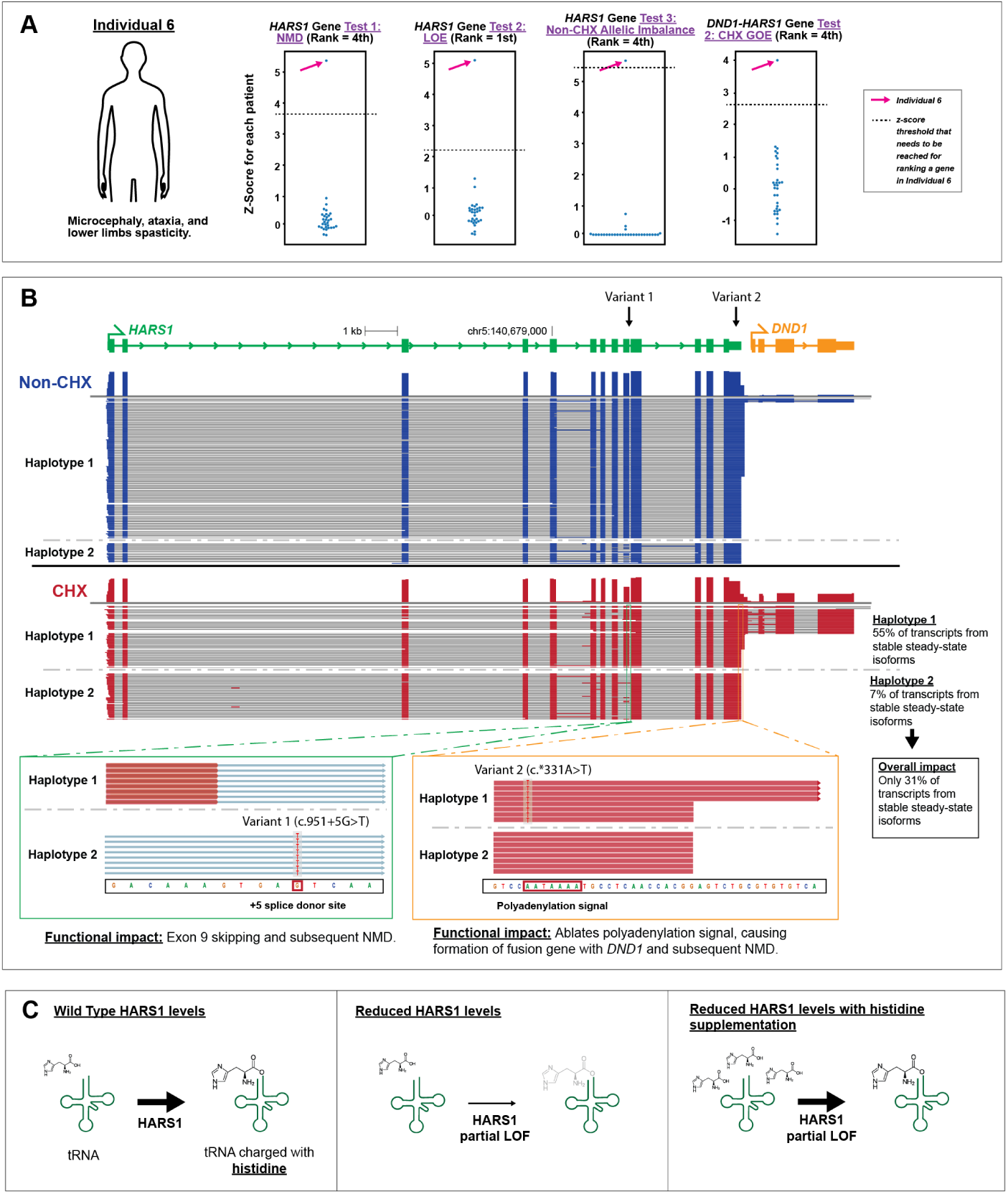
IsoRanker enables discovery of genetic cause and initiation of treatment: (A) The individual has microcephaly, ataxia, and lower limb spasticity. Z-score of the test statistic was plotted for *HARS1* across all individuals for NMD, non-treated LOE, and non-treated allelic imbalance, and for *DND1-HARS1* in CHX-treated GOE. *HARS1* ranked highly for each of these tests. (B) Genome browser view showing the reference isoform for *HARS1* (green) and neighboring gene *DND1* (orange), with observed transcripts separated by haplotype for non-treated (blue) and CHX-treated (red) samples. Two variants are indicated: Variant 1 at a splice donor site and Variant 2 at the polyadenylation signal. Zoomed views display haplotype-specific sequence reads in the CHX-treated sample, revealing Variant 1 causes exon skipping and Variant 2 causes read-through transcription. In haplotype 1 of the CHX-treated sample, 55% of the transcripts correspond to stable steady-state isoforms (i.e., isoforms that do not undergo NMD due to read-through transcription). In haplotype 2 of the CHX-treated sample, 7% of the transcripts correspond to stable steady-state isoforms (i.e., isoforms that do not undergo NMD due to exon 9 skipping). Assuming each haplotype contributes equally to total transcript output, the mean of the two haplotype percentages (31%) represents the overall level of stable transcripts. (C) Schematic of functional interpretation: under normal conditions, the HARS1 protein charges tRNA with histidine; reduced HARS1 levels cause partial loss of function (LOF), which may be partially mitigated by histidine supplementation.

Biallelic coding variants in *HARS1* have previously been associated with multisystem ataxic syndrome in three individuals.^25^ Specifically, all three individuals presented with microcephaly, intellectual disability, skeletal deformities, and ataxic broad-based gait. Individual 6 similarly presented with microcephaly, ataxia, and lower limb spasticity. Given the presence of biallelic *HARS1* variants in Individual 6, as well as their strong clinical overlap with these previously described cases, the UDN clinical team concluded that the identified *HARS1* variants were diagnostic for Individual 6’s clinical presentation. Notably, this individual had previously non-diagnostic genome, exome, and steady-state short-read RNA-seq (**Table 1**), highlighting the value of this functional long-read multi-omic data in resolving the genetic diagnosis.

### Long-read multi-ome functional data pinpoints targeted therapy

Importantly, long-read transcript data permitted us to precisely quantify the functional impact of these *HARS1* variants. Specifically, comparison of the untreated and CHX-treated samples enabled us to quantify the proportion of produced transcripts that are undergoing NMD per haplotype. For the haplotype carrying the polyadenylation signal variant, transcript usage is distributed as follows: 30% terminate using the primary polyadenylation signal and produce a canonical 3’ end. Another 25% use an alternative polyadenylation signal (AGUAAA) located 96 bp downstream and produce transcripts that are stable at steady-state. The remaining 45% do not use a polyadenylation signal within *HARS1*; instead, transcription extends into the downstream gene located 327 bp away, and termination occurs at the native polyadenylation signal of *DND1*, producing fusion isoforms containing both the spliced *HARS1* and *DND1* genes, which undergoes NMD. For the haplotype carrying the splice donor variant, 93% of transcripts undergo skipping of exon 9 and are subjected to NMD, with only 7% of transcripts retaining exon 9 and are stable at steady-state. Taking the results from both haplotypes, we can calculate that 69% of the total transcripts produced from the *HARS1* gene in Individual 6 fibroblasts are unstable at steady-state and thus unavailable for translation, with the remaining 31% of stable transcripts able to produce a full-length protein product (**Fig. 6b**).

This quantitative observation implies that *HARS1*-related multisystem ataxic syndrome is caused by a dosage-sensitive mechanism, a mechanism that has previously been shown to be responsive to supplementation in other conditions caused by aminoacyl-tRNA synthetases ^26–28^, as the enzymatic defect can be corrected via mass action. *HARS1* encodes histidyl-tRNA synthetase, an essential enzyme responsible for charging histidine onto its cognate tRNA. Consequently, this long-read multi-ome data established the basis for the clinical team to start Individual 6 on histidine supplementation (**Fig. 6c**), which has recently been explored in a clinical trial for individuals with *HARS1* deficiency (NCT02924935).

## Discussion

Our study establishes the groundwork for using a long-read transcriptome-wide approach for identifying functionally relevant non-coding variants that are causative for Mendelian conditions. Specifically, we demonstrate that pairing individual-derived full-length transcript sequencing with IsoRanker enables the individual-specific prioritization of genes and isoforms according to functional changes, such as susceptibility to NMD, expression outliers, and allelic imbalance.

An unanswered practical consideration for the broader application of individual-derived long-read transcript sequencing data is the number of individuals and sequencing depth required to achieve robust prioritization of disease-relevant transcript alterations. Our benchmarking demonstrated that IsoRanker maintained stable performance with as few as 11 individuals, suggesting that although larger cohort sizes may be important for identifying clinically relevant alterations, moderately sized rare disease cohorts can still provide sufficient ability for outlier detection. Similarly, downsampling analyses of sequencing data indicated that ∼5 million full-length non-chimeric reads per sample were sufficient to recapitulate accurate prioritization of known transcript alterations. Beyond sample size and read depth, we also observed that the choice of *de novo* isoform caller directly impacts performance, with callers that may be optimized to find the most parsimonious isoform classifications spuriously lumping rare disease-relevant transcripts with reference transcripts. Together, these findings emphasize that IsoRanker performance can be robust at practical scales of sequencing and sample collection, but careful consideration of isoform caller choice is critical for detecting the full spectrum of transcript alterations.

Furthermore, we demonstrate the utility of integrating NMD inhibition during the sample preparation step when generating long-read data for rare disease discovery purposes. Specifically, we demonstrate that NMD inhibition enables the robust identification of disease-relevant transcripts that are otherwise rarely present in steady-state transcript data, permitting the identification of disease-relevant transcripts with modest sequencing depth and thus significantly lower overall sequencing cost. We also demonstrate that NMD inhibition provides mechanistic insight into the functional effects of identified variants, which we demonstrate with Individual 6 is directly pertinent to resolving the mechanism of action of a given disease and enabling precision therapeutic decisions.

Specifically, our findings reveal that *HARS1*-related multisystem ataxic syndrome is caused by a partial loss-of-function mechanism which we anticipate is amenable to histidine supplementation. This condition has been previously reported in individuals with compound heterozygous variants^25^, with one presumed loss-of-function (LOF) allele and one missense allele of uncertain molecular consequence (i.e., it was unclear if this second missense allele was acting in a LOF, dominant-negative, or gain-of-function manner). *HARS1* is known to be dosage sensitive, as heterozygous carriers of LOF variants, who presumably have 50% residual expression, remain clinically unaffected ^25^, whereas complete loss-of-function of ARS genes is embryonically lethal in mouse models ^29^. Individual 6’s measured residual transcript level of 31% provides an important reference point, as it shows that disease manifests when *HARS1* activity falls below 50% but remains above zero. This “Goldilocks zone” of partial enzymatic function strongly suggests that the missense alleles reported in other individuals are hypomorphic, or partial LOF variants. When combined with another LOF allele, these variants reduce total activity into a range that causes disease. Notably, other conditions caused by aminoacyl-tRNA synthetase deficiencies involving partial LOF mechanisms have been shown to respond to supplementation with the precursor amino acid through a mass-action effect ^26–28^, and it was unclear if *HARS1*-related multisystem ataxic syndrome would be similarly amenable to histidine supplementation. By providing functional evidence that *HARS1*-related multisystem ataxic syndrome is caused by partial LOF, we are establishing that this condition may similarly be amenable to treatment via histidine supplementation via mass action.

Notably, although the *HARS1* splice donor variant we identified is classified as likely pathogenic and the 3’ UTR is classified as a VUS under ACMG/AMP criteria ^23,24^ (**Supplementary Table 3**), the combined clinical and functional evidence strongly supports that these *HARS1* variants represent the causative variants underlying Individual 6’s condition. Notably, as the 3’ UTR variant results in only a partial LOF (i.e., only 45% of transcripts are subjected to NMD), it is anticipated that individuals homozygous for this variant would not develop a *HARS1*-related phenotype. As such, this variant could be considered conditionally pathogenic only when present in *trans* with a more severe *HARS1* LOF variant, highlighting a deficiency of the current ACMG classifications for variants with partial LOF in autosomal recessive conditions caused by a dosage-sensitive LOF mechanism.

While IsoRanker successfully prioritized all five known transcript-altering events within the top 20 ranked genes or isoforms in the individuals and tests of interest, the highest-ranked signal is not necessarily the causal one. On average, each individual carries approximately 100 loss-of-function variants that can trigger NMD, many of which occur in non-essential genes and are well tolerated.^30^ As a result, some genes and isoforms may rank highly in our transcriptome-based analysis due to genuine functional alterations that are incidental to the individual’s disease. Therefore, careful interpretation of IsoRanker outputs in the context of biological plausibility, such as phenotype concordance, is essential for distinguishing truly pathogenic signals.

Overall, our study provides further evidence for the utility of augmenting genomic sequencing with function-based approaches for identifying pathogenic causes of rare diseases. Furthermore, our study highlights the value of accomplishing this using long-read transcript sequencing, and by pairing steady-state measures of transcript abundance with those obtained after treating cells with an NMD inhibitor. Transcript alterations are only one of the many non-coding functional alterations that can cause disease ^31^, and moreover, many transcripts may be expressed in cells or at developmental stages that are not readily obtained clinically. Consequently, we anticipate that the utilization of additional *in vitro* derived cell types, as well as the use of multi-omic approaches that integrate additional functional modalities^10^ will expand our ability to identify non-coding variants that are contributing to rare diseases.

## Data Availability

Sequencing data have been deposited in NHGRI's Analysis Visualization and Informatics Lab-space (AnVIL) under accession number phs003047. The Undiagnosed Diseases Network (UDN) data can be identified by the internal_project_id field containing "pnw-udn-long-read-multi-ome". Data from the Manton cohort are labeled using the Broad_datatype_ID as part of the sample identifier; for example, Broad_genome_MAN_1877-01_D1_1 represents one of the deposited samples in AnVIL.

## Supplementary Figures

**Supplementary Figure 1.**
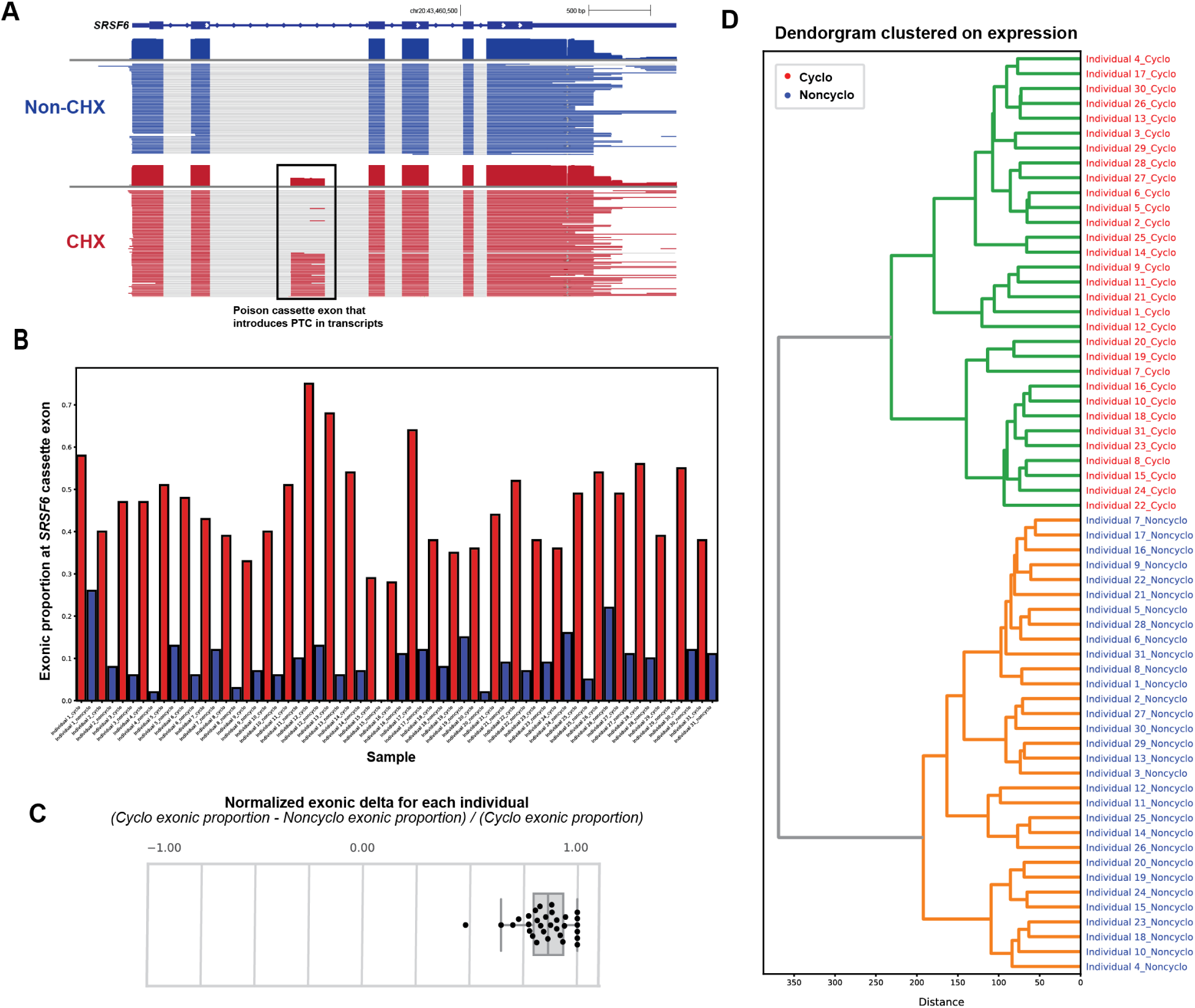
Validating effects of cycloheximide: (A) *SRSF6* harbors a poison cassette exon that introduces a premature termination codon (PTC). When this exon is included in the transcript, the transcript is degraded via NMD during steady-state. Thus, detection of the cassette exon when a sample is treated with CHX is useful for validating the effects of CHX on inhibiting NMD. (B) For each individual, the proportion of *SRSF6* reads with inclusion of the cassette exon was calculated and plotted for the CHX-treated (red) sample and the untreated sample (blue). In each individual, the CHX-treated sample had much greater proportion of reads with the NMD-inducing cassette exon. This validated the effect of CHX on inhibiting NMD for each individual. (C) The normalized exonic delta at the position of the *SRSF6* cassette exon for each individual was calculated and plotted. (D) Gene-level expression matrix was used for hierarchical clustering and generation of a dendrogram.

**Supplementary Figure 2.**
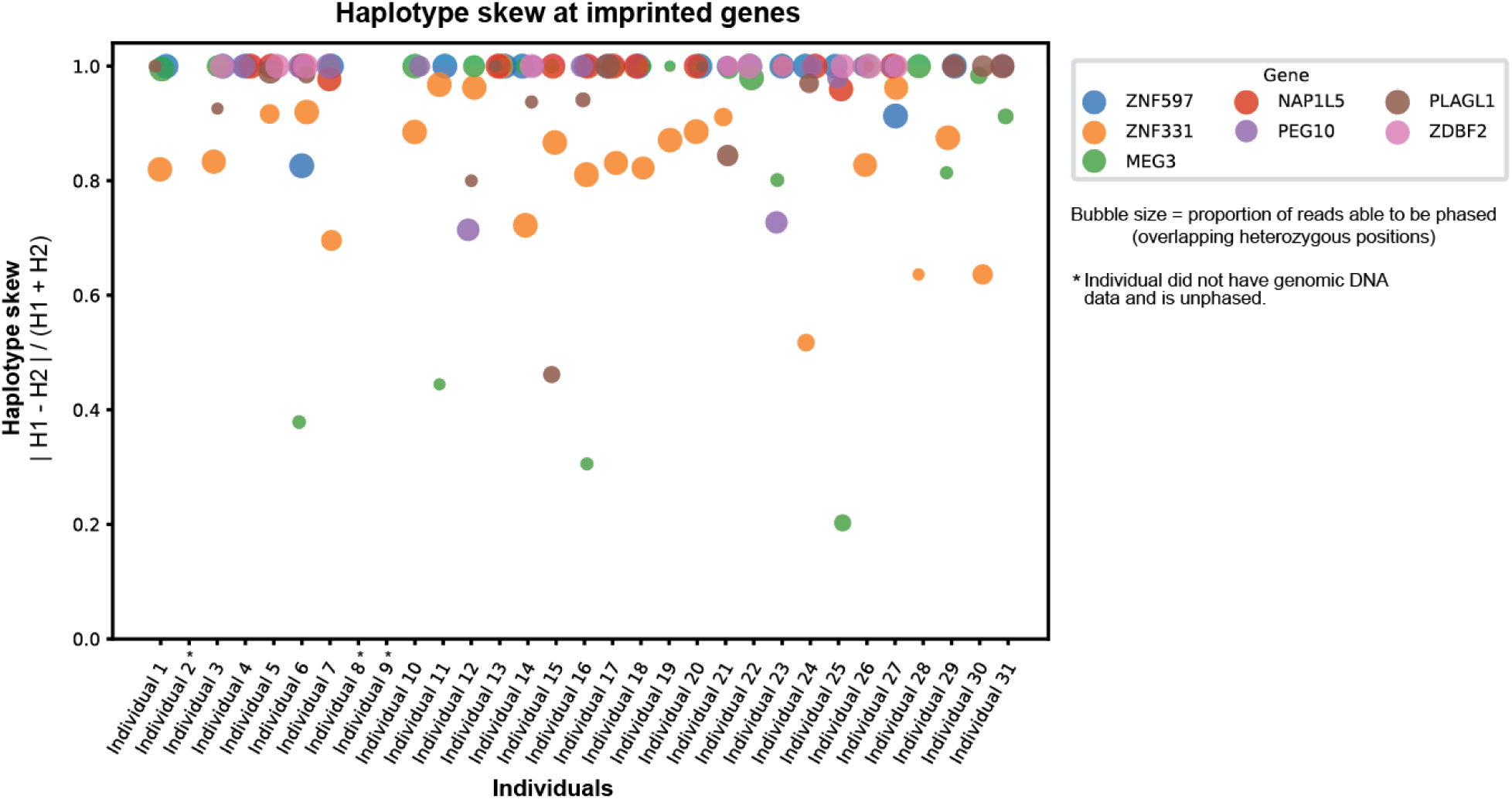
Validating ability to phase transcript data: Seven genes known to be imprinted and expressed in fibroblasts were selected to be used to evaluate our ability to appropriately phase transcript data. The majority of expression from these genes showed strong haplotype skewing, indicating successful phasing of transcripts.

**Supplementary Figure 3.**
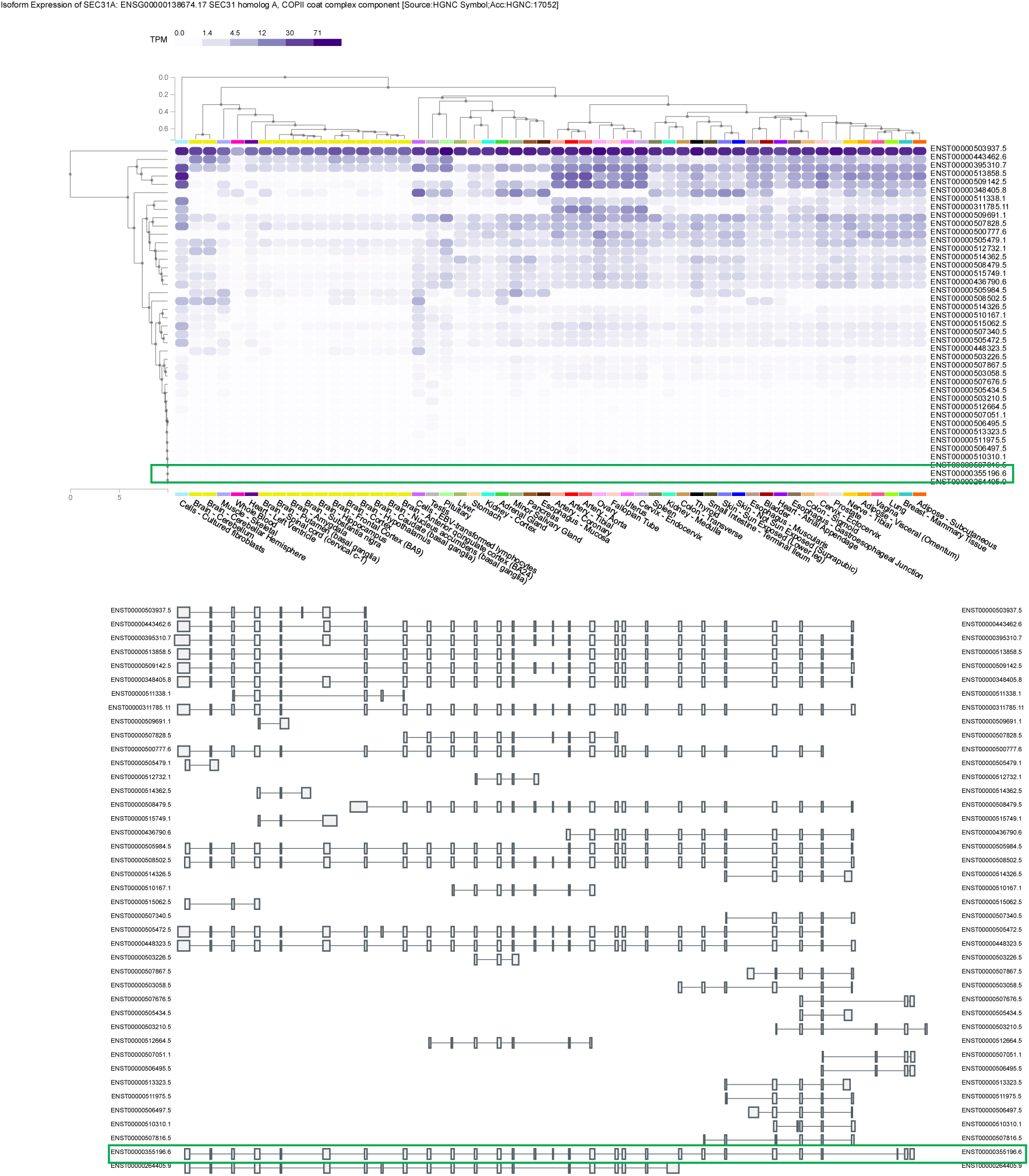
*SEC31A* GTEx isoform expression: The *SEC31A* isoform (ENST00000355196.6) that overlaps the splice donor variant in Individual 5 is a minimally expressed isoform in all tissues.

## Supplementary Tables

**Supplementary Table 1.**
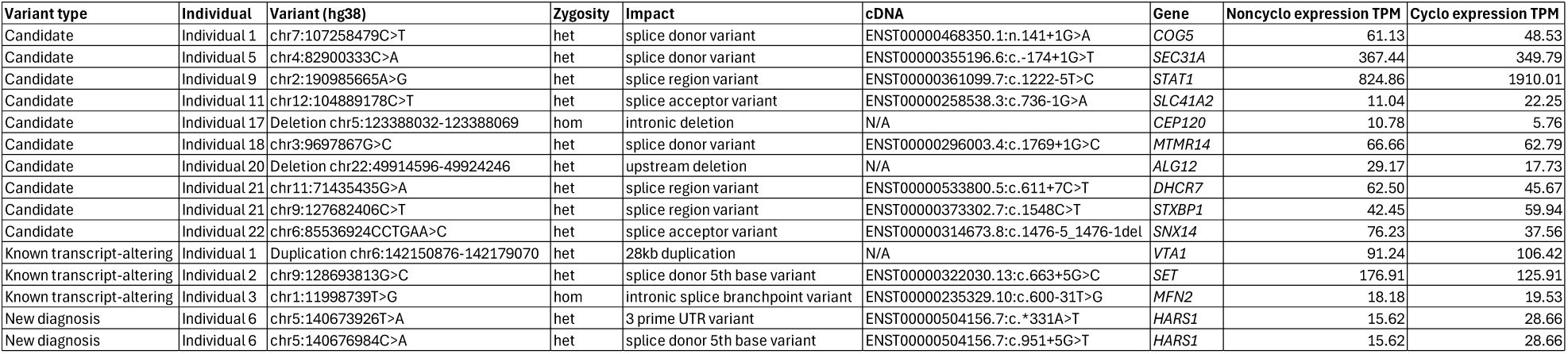
Details of genetic variants: The table summarizes the specific variants highlighted in the manuscript, including their type, genomic coordinates (hg38), zygosity, predicted functional impact, and affected isoform (cDNA) and gene. For each individual carrying the variant, gene expression levels (in transcripts per million, TPM) are shown under noncycloheximide (Noncyclo) and cycloheximide (Cyclo) treatment conditions.

**Supplementary Table 2.**
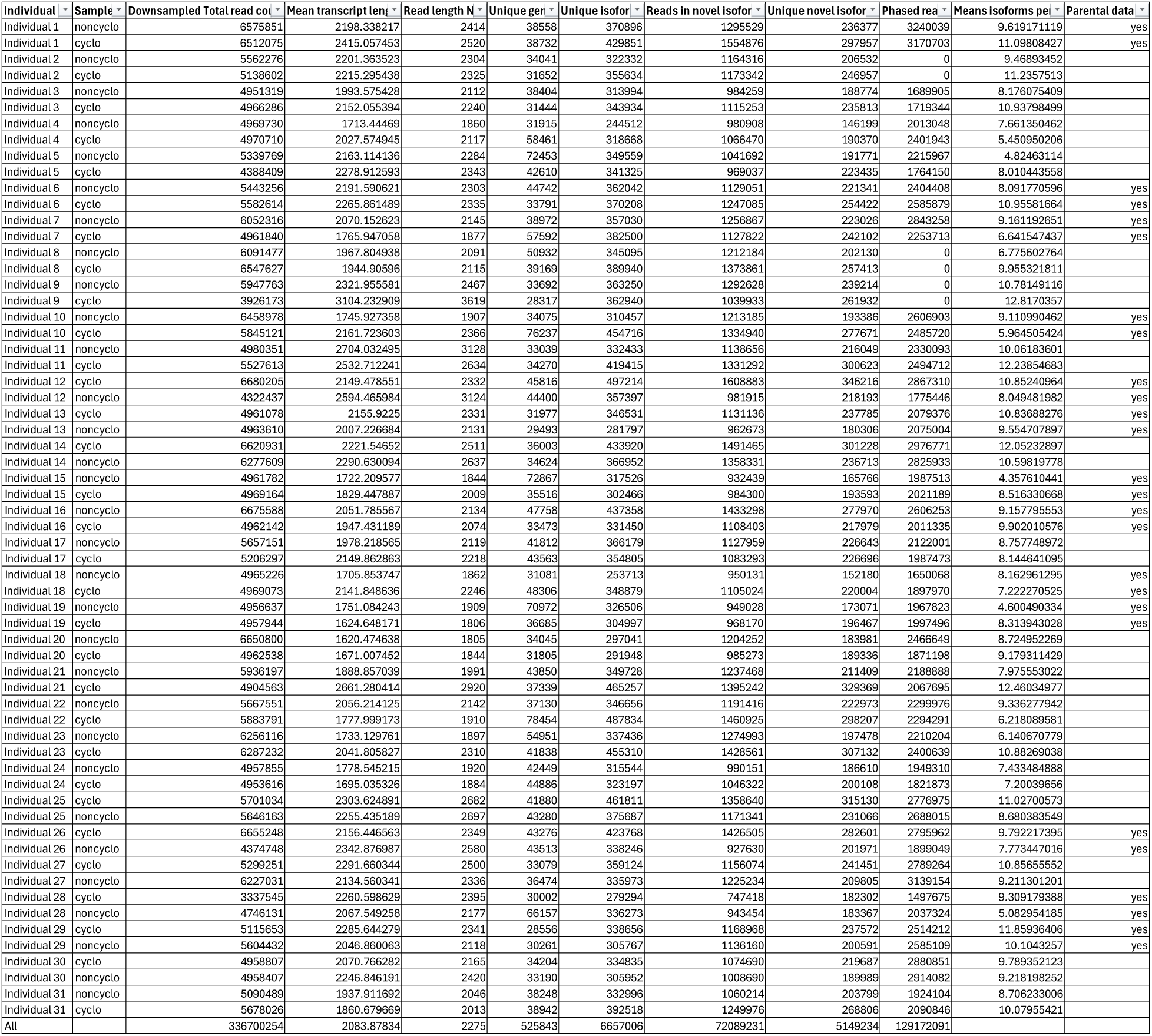
Summary of quality control metrics for long-read transcriptome sequencing data: The table lists, for each individual and sample type, the total number of downsampled reads, mean transcript length, read length N50, number of unique genes and isoforms detected, proportion of reads assigned to novel isoforms, number of unique novel isoforms, number of phased reads, mean isoforms per gene, and availability of corresponding parental data.

**Supplementary Table 3.** Interpretation of Individual 6 *HARS1* variants using ACMG guidelines.

| EVIDENCE FOR PATHOGENICITY: |  |  |  |  |
| --- | --- | --- | --- | --- |
| RULE | STRENGTH | BRIEF DESCRIPTION | chr5:140676984C>A<br>(NM_002109.6:c.951+5G>T) | chr5:g.140673926T>A<br>(NM_002109.6:c.*331A>T) |
| PVS1 | Very Strong | Null variant (w/ LOF mechanism) | PVS1 |  |
| PS1 | Strong | Different nt change (same aa) as a path variant |  |  |
| PS2 | Strong | *De novo* (mat/pat confirmed) |  |  |
| PS3 | Strong | Established functional data shows deleterious |  | PS3_supporting |
| PS4 | Strong | Case data or case-control studies |  |  |
| PM1 | Moderate | Mutation hotspot/critical functional domain |  |  |
| PM3 | Moderate | *In trans* with path variant (AR) |  | PM3 |
| PM4 | Moderate | In-frame indels, stop-loss variants |  |  |
| PM5 | Moderate | Different aa change at codon is path |  |  |
| PM6 | Moderate | Assumed *de novo* (no mat/pat testing) |  |  |
| PM2 | Supporting | Absent/rare in controls | PM2_supporting | PM2_supporting |
| PP1 | Supporting | Segregation |  |  |
| PP2 | Supporting | Missense in a gene with low rate of variation |  |  |
| PP3 | Supporting | Computational evidence predicts impact |  |  |
| PP4 | Supporting | Disease with single genetic etiology |  |  |
| PP5 | Supporting | Reputable source reports variant as path |  |  |
| N/A | Any strength | Other evidence not captured by ACMG |  |  |
| EVIDENCE FOR BENIGN: |  |  |  |  |
| RULE | STRENGTH | BRIEF DESCRIPTION | Applied? | Applied? |
| BA1 | Stand Alone | >5% MAF |  |  |
| BS1 | Strong | MAF greater than disorder frequency |  |  |
| BS2 | Strong | Observed in healthy individual |  |  |
| BS3 | Strong | Established functional data shows no effect |  |  |
| BS4 | Strong | Lack of segregation |  |  |
| BP1 | Supporting | Missense in gene with only truncating mutations |  |  |
| BP2 | Supporting | *In trans* (AD) or *in cis* (all) with path variant |  |  |
| BP3 | Supporting | In-frame indels in repetitive region |  |  |
| BP4 | Supporting | Computation evidence predicts no impact |  | BP4 |
| BP5 | Supporting | Case has alternate molecular basis for disease |  |  |
| BP6 | Supporting | Reputable source reports variant as benign |  |  |
| BP7 | Supporting | Synonymous with no splice impact/conservation |  |  |
| N/A | Any strength | Other evidence not captured by ACMG |  |  |
|  |  | Classification submitted to ClinVar: | Likely Pathogenic | Uncertain Significance |

**Supplementary Table 4.** Lookup table for external IDs.

| Individual | External ID |
| --- | --- |
| Individual 1 | UDN687128 |
| Individual 2 | UDN215640 |
| Individual 3 | UDN633333 |
| Individual 4 | UDN318336 |
| Individual 5 | UDN052264 |
| Individual 6 | UDN212054 |
| Individual 7 | UDN355246 |
| Individual 8 | UDN860070 |
| Individual 9 | UDN353099 |
| Individual 10 | UDN305049 |
| Individual 11 | UDN310876 |
| Individual 12 | MAN_1199-01 |
| Individual 13 | MAN_1877-01 |
| Individual 14 | MAN_310-01 |
| Individual 15 | UDN121217 |
| Individual 16 | UDN374570 |
| Individual 17 | UDN827597 |
| Individual 18 | UDN966087 |
| Individual 19 | UDN204349 |
| Individual 20 | UDN359892 |
| Individual 21 | UDN262407 |
| Individual 22 | UDN924176 |
| Individual 23 | UDN666440 |
| Individual 24 | UDN178694 |
| Individual 25 | MAN_1986-01 |
| Individual 26 | MAN_0252-01 |
| Individual 27 | MAN_2275-01 |
| Individual 28 | MAN_2401-01 |
| Individual 29 | MAN_2410-01 |
| Individual 30 | MAN_2443-01 |
| Individual 31 | UDN010451 |

## Acknowledgements

We thank the research participants and their families in the UDN and GREGoR cohorts. We also thank the UW PacBio sequencing center for their assistance in performing the transcriptome and long-read sequencing and Elizabeth Tseng for her guidance with processing the long-read transcript data. We thank Jill Madden for assisting in obtaining samples for this project, as well as the UDN clinical investigators. Research reported in this publication was supported by the National Institute of Neurological Disorders and Stroke of the National Institutes of Health under Award Number(s) U01HG010233 and U01NS134355, as well as a pilot award from the UDN. Sequence data for the MAN samples was provided by the Manton Center for Orphan Disease Research Pilot Award to A.B.S. and A.O’D-L.. A.B.S. holds a Career Award for Medical Scientists from the Burroughs Wellcome Fund and is a Pew Biomedical Scholar. This study was supported by National Institutes of Health (NIH) grants 1DP5OD029630, and 1U01HG013744 to A.B.S.. This was also supported by CZIF2024-010284 from the Chan Zuckerberg Initiative (CZI). M.R.V. was supported by a training grant (T32) from the NIH (2T32GM007454-46) and a Pathway to Independence award from the National Institute of General Medical Sciences (1K99GM155552-01). A.O’D-L. was supported by U01HG011755. Sequence data analysis was supported by the University of Washington Center for Rare Disease Research (UW-CRDR), which is funded by NHGRI grant U01 HG011744. The content is solely the responsibility of the authors and does not necessarily represent the official views of the National Institutes of Health.

## Author contributions

Y.H.H.C., A.O’D-L. and A.B.S. conceived and designed the study. Y.H.H.C. developed the IsoRanker computational framework and performed all data analyses. Y.H.H.C. and A.B.S. wrote the manuscript. A.B.S. supervised the study and provided conceptual guidance throughout. Y.H.H.C, J.E.R, and K.M.M. assisted with long-read sequencing data generation and interpretation. A.E.S developed IsoSeq_smk and IsoRanker_vis for processing the long-read sequencing data. D.M.W., M.R.V, and A.E.S. provided statistical and computational advice for framework development. C.A.G., and E.B. provided clinical support. E.E.B, R.D.K., and J.X.C provided evaluation of the genetic data. M.H.W., A.H.B, M.J.B, C-L.W., K.M.D., G.J., A.O’D-L., and A.B.S. provided funding. The PNW-UDN and UW and Broad GREGoR sites contributed clinical and genetic expertise and facilitated access to individual samples. The UW-CRDR contributed genetic expertise. All authors reviewed and approved the final manuscript.

## Data and code availability

Sequencing data have been deposited in NHGRI’s Analysis Visualization and Informatics Lab-space (AnVIL) under accession number phs003047. The Undiagnosed Diseases Network (UDN) data can be identified by the internal_project_id field containing “pnw-udn-long-read-multi-ome”. Data from the Manton cohort are labeled using the Broad_datatype_ID as part of the sample identifier; for example, Broad_genome_MAN_1877-01_D1_1 represents one of the deposited samples in AnVIL. All code used in this study is available at: https://github.com/yhhc2/IsoRanker/tree/main/Paper.

## Consortia

Members of the Undiagnosed Diseases Network (UDN): Please see UDN banner authorship document.

Members of the Genomics Research to Elucidate the Genetics of Rare Diseases consortium (GREGoR): Please see GREGoR banner authorship document.

Members of the University of Washington Center for Rare Disease Research (UW-CRDR): Please see UW-CRDR banner authorship document.

